# Altered brain energy metabolism related to astrocytes in Alzheimer’s disease

**DOI:** 10.1101/2023.03.09.23286881

**Authors:** Kosei Hirata, Kiwamu Matsuoka, Kenji Tagai, Hironobu Endo, Harutsugu Tatebe, Maiko Ono, Naomi Kokubo, Asaka Oyama, Hitoshi Shinotoh, Keisuke Takahata, Takayuki Obata, Masoumeh Dehghani, Jamie Near, Kazunori Kawamura, Ming-Rong Zhang, Hitoshi Shimada, Takanori Yokota, Takahiko Tokuda, Makoto Higuchi, Yuhei Takado

## Abstract

**Objective:** Increasing evidence suggests that reactive astrocytes are associated with Alzheimer’s disease (AD). However, its underlying pathogenesis remains unknown. Given the role of astrocytes in energy metabolism, reactive astrocytes may contribute to altered energy metabolism. It is hypothesized that lactate, a glucose metabolite, is produced in astrocytes and subsequently shuttled to neurons as an energy substrate. This study aimed to examine alterations in brain lactate levels and their association with astrocytic activities in AD.

**Methods:** 30 AD and 30 cognitively unimpaired (CU) subjects were enrolled. For AD subjects, amyloid and tau depositions were confirmed by positron emission tomography using [^11^C]PiB and [^18^F]florzolotau, respectively. Lactate and myo-inositol, an astroglial marker, in the posterior cingulate cortex (PCC) were quantified by magnetic resonance spectroscopy (MRS). These MRS metabolites were compared with plasma biomarkers, including glial fibrillary acidic protein (GFAP) as another astrocytic marker.

**Results:** Lactate and myo-inositol levels were higher in AD than in CU (*p* < 0.05). Lactate levels correlated with myo-inositol levels (*r* = 0.272, *p* = 0.047). Lactate and myo-inositol levels were positively associated with the Clinical Dementia Rating sum-of-boxes scores (*p* < 0.05). Significant correlations were noted between myo-inositol levels and plasma GFAP and tau phosphorylated at threonine 181 levels (*p* < 0.05).

**Interpretation:** We found high lactate levels accompanied by an increased astrocytic marker in the PCC in AD. Thus, impaired lactate shuttle of reactive astrocytes may disrupt energy regulation, resulting in surplus lactate levels. Myo-inositol and plasma GFAP may reflect similar astrocytic changes.

## INTRODUCTION

Alzheimer’s disease (AD) is the most common form of dementia and is currently defined based on the deposition of amyloid beta (Aβ) and hyperphosphorylated tau protein (p-tau) in the brain.^1^ Recently, the role of glial cells in the neurodegenerative process of AD has received more attention.^2^ Astrocytes, which are responsible for maintaining central nervous system homeostasis and supporting neuronal function, undergo structural and biochemical changes in response to pathological conditions with various effects on the pathophysiology.^3, 4^ Although reactive astrocytes have been implicated in AD initiation and progression,^2^ how these reactive astrocytes contribute to neurodegeneration remains unknown.

Appropriate regulation of brain energy metabolism is crucial for the efficient functioning of the central nervous system.^5^ Emerging evidence suggests that metabolic changes in the brain are associated with AD pathophysiology, as demonstrated by previous positron emission tomography (PET) studies using 2-[^18^F]fluoro-2-deoxy-D-glucose.^6^ Given the role of astrocytes in providing metabolic support to neurons,^5, 7^ astrocyte reactivity may be important in understanding the underlying mechanisms of energy metabolic changes in AD. Regarding astrocyte–neuron metabolic coupling, lactate may be a key molecule, as the astrocyte–neuron–lactate shuttle (ANLS) model has been proposed.^8^

According to this model, lactate produced through glycolysis in astrocytes is supplied to neurons and contributes to neuronal energy production. The ANLS is essential for long-term memory formation, and a recent study found an association between high brain lactate levels and cognitive decline in mouse models of neuropsychiatric diseases including AD.^9-11^ These findings suggest that lactate is not merely a metabolic waste product but can be a vital energy molecule produced by astrocytes. However, whether and how lactate concentrations in the brain are altered in patients with AD is not well known. We hypothesized that reactive astrocytes contribute to altered brain energy metabolism in AD pathophysiology.

Proton magnetic resonance spectroscopy (^1^H-MRS) allows for the non-invasive measurement of tissue metabolites, providing insights into specific cellular and molecular changes in the brain. Myo-inositol, which is widely accepted as an astrocytic marker, is high in patients with AD.^12^ Lactate can be measured by MRS; however, measuring lactate levels by conventional MRS is difficult because of its low concentration in the normal brain. The only MRS study that evaluated lactate levels in patients with AD found high lactate levels in the posterior cingulate cortex (PCC), although the authors acknowledged that their MRS sequence had technical limitations for lactate quantification.^13^ Therefore, to capture subtle metabolite changes, the present study used an advanced MRS sequence recommended by an expert consensus^14^ and a head coil with a higher number of channels than that used in the previous study^13^ to assess lactate levels and their association with myo-inositol levels in the brains of patients with AD. Furthermore, by examining the correlation of these MRS metabolites with cognitive function scores and plasma biomarkers, including another astrocytic marker (glial fibrillary acidic protein [GFAP]),^15^ Aβ (Aβ 42 and Aβ 40), and tau (p-tau 181), this study aimed to provide novel insights into the role of reactive astrocytes in AD pathogenesis.

## MATERIALS AND METHODS

### Participants

Participants with AD were prospectively recruited from the affiliated hospitals of the National Institute of Radiological Sciences (NIRS) and National Institutes for Quantum Science and Technology (QST) between November 2018 and February 2022. The AD group included cases with mild cognitive impairment (MCI) due to AD and AD dementia, all of which met Petersen’s criteria^16^ and NINDS-ADRDA criteria,^17^ respectively. Age- and sex-matched cognitively unimpaired participants (CU group) were recruited from the volunteer association of the NIRS-QST and did not have a history of neurologic or psychiatric disorders. At the screening, all participants with AD were confirmed to be amyloid-positive based on visual assessment of PET images of [^11^C]Pittsburgh Compound-B (PiB) by three readers according to the established criteria.^18^ In addition, AD diagnosis with high probability was achieved through the assessment of tau deposition by tau PET using [^18^F]florzolotau.^19^ AD severity was assessed using the Clinical Dementia Rating (CDR) and CDR Sum-of-Boxes (CDR-SOB) score, which ranges from 0 to 18,^20^ and all participants were evaluated for cognitive function using the Mini-Mental State Examination (MMSE).^21^ Finally, a total of 30 patients with AD and 30 CU individuals were enrolled in the study.

This study was approved by the Institutional Review Board of QST. Written informed consent was obtained from all participants and/or from spouses or other close family members of participants who were cognitively impaired.

### Magnetic resonance imaging (MRI)/MRS data acquisition

MRI and MRS were conducted using a 3.0-T scanner (MAGNETOM Verio; Siemens Healthcare) equipped with a 32-channel receiving head coil. Structural three-dimensional T1-weighted images were acquired by magnetization-prepared rapid gradient-echo sequence (MPRAGE, repetition time [TR] = 2,300 ms; echo time [TE] = 1.95 ms; TI = 900 ms; field of view [FOV] = 250 mm; flip angle = 9°; acquisition matrix = 512 × 512; and axial slice thickness, 1 mm). Single-voxel MRS data were acquired using a short TE spin-echo full-intensity acquired localized single voxel spectroscopy (SPECIAL) sequence^22^ with TE of 8.5 ms, TR of 3000 ms, and 128 averages as described previously.^23^ The SPECIAL spectroscopy sequence was specifically developed to obtain a full-intensity signal in a defined volume of interest (VOI) using an ultra-short TE. The VOI was placed at the PCC with the sizes 20 × 20 × 20 mm^3^ (Figure 1A). This region has been demonstrated to have decreased glucose utilization in AD^6^ and has been recommended by the MRS Consensus Group for MRS research in AD.^24^

**Figure 1.**
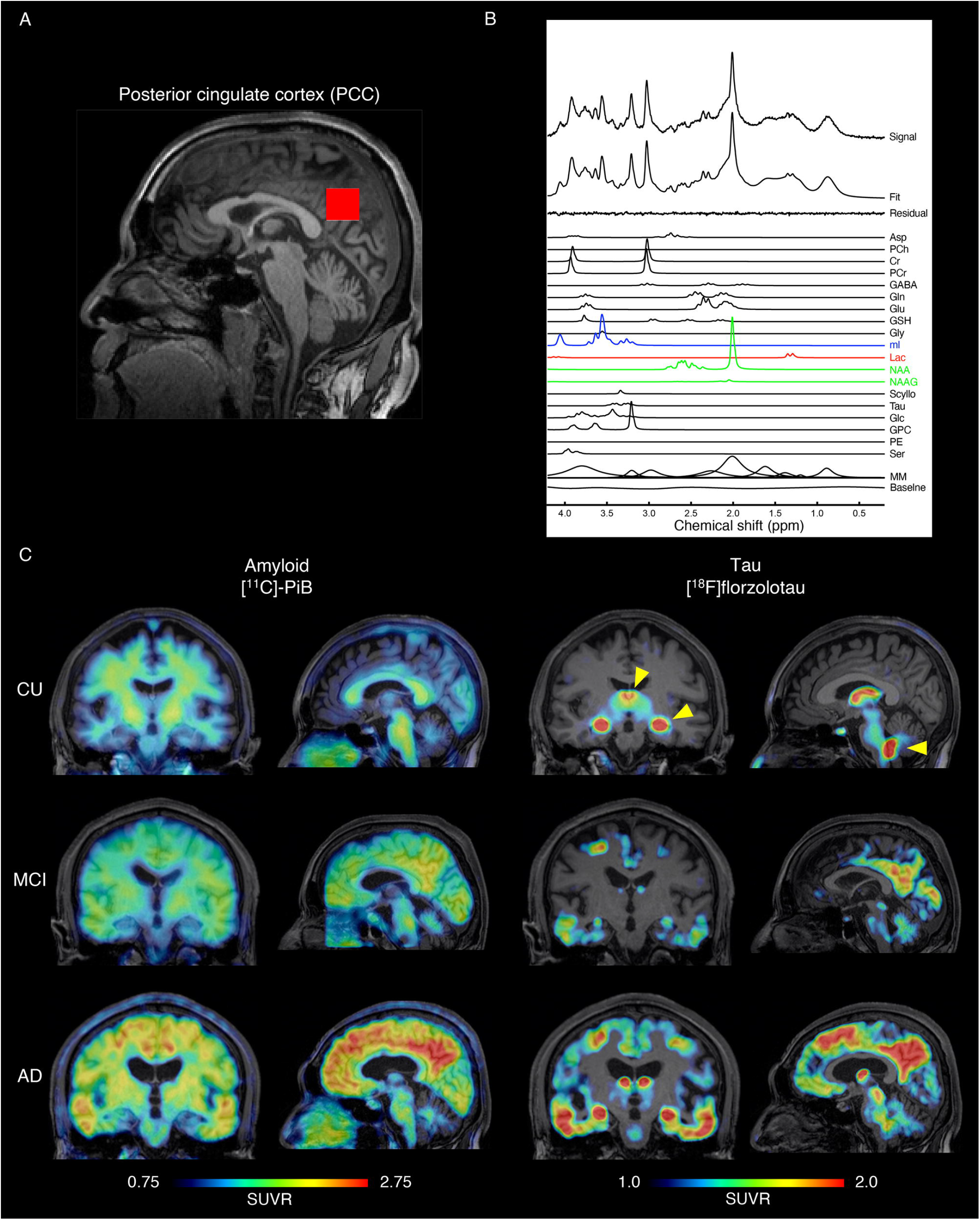
Voxel placement, example spectra, and representative amyloid and tau PET images. (A) A representative single-voxel placed in the PCC with voxel volumes 20 × 20 × 20 mm^3^ overlaid onto a T1-weighted image. (B) An example MRS spectrum in a patient with AD obtained from the voxel. MRS data were acquired with SPECIAL sequence and analyzed by LCModel. LCModel spectral fit, fit residual, macromolecules (MM), baseline, and individual metabolite fits including myo-inositol (blue), lactate (red), and tNAA (green) are shown. (C) Representative images of [^11^C]PiB amyloid PET and [^18^F]florzolotau tau PET images across diagnostic groups. Yellow arrowheads indicate nonspecific binding in the choroid plexus. AD = Alzheimer disease; CU = cognitively unimpaired controls; MCI = mild cognitive impariment; MRS = magnetic resonance spectroscopy; PCC = posterior cingulate cortex; SPECIAL sequence = short TE spin-echo full-intensity acquired localized single voxel spectroscopy sequence; SUVR = standardized uptake value ratio; tNAA = total N-acetyl-aspartate.

### MRS analysis

One AD patient was unable to tolerate the MRI scan and was excluded from further analysis. Before signal averaging and data analysis, a weighted combination of receiver channels was processed using the FID-A toolkit.^25^ The removal of motion-corrupted averages, spectral registration for frequency and phase drift correction, and alignment of subspectra before subtraction were performed. MRS data were analyzed using LCModel software (Stephen Provencher, Inc.) for a linear combination of model spectra with simulated basis functions.^26^ The neurochemical basis set used included nine macromolecular basis functions, each derived from the individual peaks in a macromolecular spectrum that was obtained by summing experimentally acquired metabolite-nulled spectra from six healthy adult volunteers. An example spectrum is shown in Figure 1B. The spectral signal-to-noise ratio (SNR) and linewidth (LCModel output) had average ± standard deviation (SD) of 68.5 ± 12.6 and 3.962 ± 0.767 Hz, respectively. All MRS data were confirmed to meet the quality criteria for the SNR and linewidth quality criteria.^27^ In this study, we assessed the levels of myo-inositol, lactate, and total N-acetyl-aspartate (tNAA: NAA + N-acetyl–aspartyl–glutamate [NAAG]) as a neuronal marker. One participant with AD and two CU individuals were excluded for lactate assessment due to the presence of severe artifacts, resulting in a lack of output data. Neurochemical quantification reliability was evaluated using the Cramer-Rao lower bound (CRLB) provided by LCModel on a per-metabolite basis. Our study employed a cutoff of 30% CRLB averaged across all scans.

Excluding individual samples with high CRLB values, particularly for metabolites with low concentrations, is not recommended, as it can introduce bias in the mean concentrations of the entire cohort.^28^ The average ± SD of CRLB for myo-inositol, lactate, and tNAA were 3.5% ± 0.7%, 24.9% ± 11.1%, and 2.0% ± 0.4%, respectively, all of which met the criteria. Concentrations were quantified using tissue water as an internal reference. Partial volume effects were corrected by extracting the fractions of the gray matter (GM), white matter (WM), and cerebrospinal fluid (CSF) within the VOIs calculated via the segmentation of T1-weighted images using Gannet 3.0 software.^29^ Water concentrations of GM, WM, and CSF were assumed to be 35,880, 43,300, and 55,556 mM, respectively. Water concentrations were then corrected based on the partial volume fractions of the GM, WM, and CSF using the equation previously described.^30^ Metabolite concentrations (μmol/g) were divided by the GM and WM fractions to correct for CSF in the VOI.

### PET acquisition and analysis

Cerebral Aβ and tau depositions were examined using [^11^C]PiB and [^18^F]florzolotau PET, respectively. [^11^C]PiB and [^18^F]florzolotau were radiosynthesized according to the protocols of our previous studies.^19^ The participants were injected intravenously with approximately 185 MBq of ^1^[^18^F]florzolotau and scanned for 90–110 min after injection. All images were collected using the Biograph mCT Flow system (Siemens Healthcare). Meanwhile, the [^11^C]PiB (injected dose: approximately 555 MBq) PET was performed, and participants were scanned for 50–70 min after injection using ECAT EXACT HR+ Scanner (CTI PET Systems, Inc.), Biograph mCT Flow system (Siemens Healthcare), or Discovery MI (GE Healthcare). All PET images were reconstructed with a filtered back projection. Images were analyzed using PMOD® software version 3.8 (PMOD Technologies Ltd., Zurich, Switzerland). Motion correction and coregistration to the corresponding T1-weighted images of the participants were performed. Standardized uptake value ratio (SUVR) images were created with the cerebellar GM as the reference region. For the reference region, surface-based cortical reconstruction and volumetric subcortical segmentation of T1-weighted images were performed using FreeSurfer software (version 6.0.0; http://surfer.nmr.harvard.edu) as described in our previous study.^23^ Representative PET images of [^11^C]PiB and [^18^F]florzolotau are shown in Figure 1C.

### Immunoassay protocols

The plasma levels of GFAP, Aβ40, Aβ42, and tau phosphorylated at threonine 181 (p-tau181) were assessed in blood samples obtained from 28 CU individuals and 27 patients with AD. These samples were analyzed by an HD-X Simoa analyzer using commercially available kits from a single lot, according to the instructions of the manufacturer (Quanterix, Lexington, MA, USA).^31^ All samples were analyzed in duplicate on a single occasion.

### Statistical analysis

Intergroup differences were evaluated using independent-samples t test for parametric data or Mann-Whitney test for non-parametric data. Partial correlation analysis, adjusted for age and sex, was used to evaluate correlations between clinical characteristics, metabolite levels, and plasma biomarkers. All statistical tests were two-tailed, and p-values of <0.05 indicated statistical significance; multiple comparisons were corrected by false discovery rate correction using Benjamini-Krieger-Yekutieli method.^32^ Statistical analyses were performed using IBM SPSS Statistics version 27 (IBM Corp., Armonk, NY, USA), with only the correction for multiple comparisons carried out using GraphPad Prism 9 software (Graph pad software, San Diego, CA, USA).

## RESULTS

### Demographic and clinical data

The demographic and clinical features of the participants are summarized in Table 1. The study population consisted of 30 CU individuals and 30 patients with AD (MCI, n = 9; AD dementia, n = 21). The two groups did not differ in age or sex. The AD group showed worse cognitive function, as demonstrated by lower total scores on the MMSE, than the CU group (*p* < 0.001). All AD patients were confirmed to have the characteristic distribution of amyloid and tau pathology through visual assessment of [^11^C]PiB and [^18^F]PMPBB3 PET imaging, respectively. Conversely, all CU participants had negative results on both amyloid and tau PET scans.

**Table 1.** Baseline characteristics

| Characteristic | CU, n = 30 | AD, n = 30 | *p* |
| --- | --- | --- | --- |
| Sex, female, n (%) | 17 (56.7) | 18 (60.0) | 0.79 |
| Age, years (SD) | 66.6 (10.1) | 68.6 (11.4) | 0.48 |
| MMSE (SD) | 29.1 (1.1) | 21.3 (3.9) | < 0.001 |
| CDR global score, n (%) |  |  |  |
| 0.5 | N/A | 9 (30) |  |
| 1 | N/A | 16 (53.3) |  |
| 2 | N/A | 5 (16.7) |  |
| CDR-SOB (SD) | N/A | 4.5 (2.8) |  |
| Myo-inositol, $\mu\text{mol/g}$ | 8.2 (1.0) | 10.2 (1.9) | <0.001<sup>a</sup> |
| Lactate, $\mu\text{mol/g}$ | 0.8 (0.4) | 1.0 (0.4) | 0.016<sup>a</sup> |
| tNAA, $\mu\text{mol/g}$ | 12.1 (1.1) | 12.1 (1.8) | 0.186<sup>a</sup> |
| Plasma GFAP, pg/mL (SD) | 105.5 (41.9) | 231.8 (107.4) | <0.001<sup>a</sup> |
| Plasma Aβ42/40 ratio (SD) | 0.092 (0.015) | 0.070 (0.011) | <0.001<sup>a</sup> |
| Plasma p-tau181, pg/mL (SD) | 1.6 (0.7) | 4.3 (1.4) | <0.001<sup>a</sup> |

Data are mean (standard deviation). AD = Alzheimer disease; Aβ 42 = amyloid $\beta$ 42; Aβ 40 = amyloid $\beta$ 40; Aβ 42/40
ratio = ratio Aβ 42 to Aβ 40; CDR = Clinical Dementia Rating; CDR SOB = Clinical Dementia Rating Sum of Boxes; CU
= cognitively unimpaired controls; GFAP = glial fibrillary acidic protein; MMSE = Mini-Mental Status Examination; SD
= standard deviation; tNAA = total N-acetyl-aspartate; p-tau181 = tau phosphorylated at threonine 181.
<sup>a</sup>False discovery rate-adjusted p value.

### MRS results

MRS metabolites measured in the PCC were evaluated for myo-inositol, lactate, and tNAA (Table 1 and Figure 2A). Myo-inositol levels were significantly high in the AD group compared with the CU group (*p* < 0.001). Lactate levels were significantly different between the AD and CU groups, with the AD group showing high lactate levels (*p* = 0.016). No significant difference in the tNAA levels was found between the two groups (*p* = 0.186). Then, whether astrocytic changes are associated with high lactate levels was evaluated by examining the relationship between myo-inositol and lactate levels. High myo-inositol levels correlated with higher lactate levels (*r* = 0.272, *p* = 0.047) (Figure 2B).

**Figure 2.**
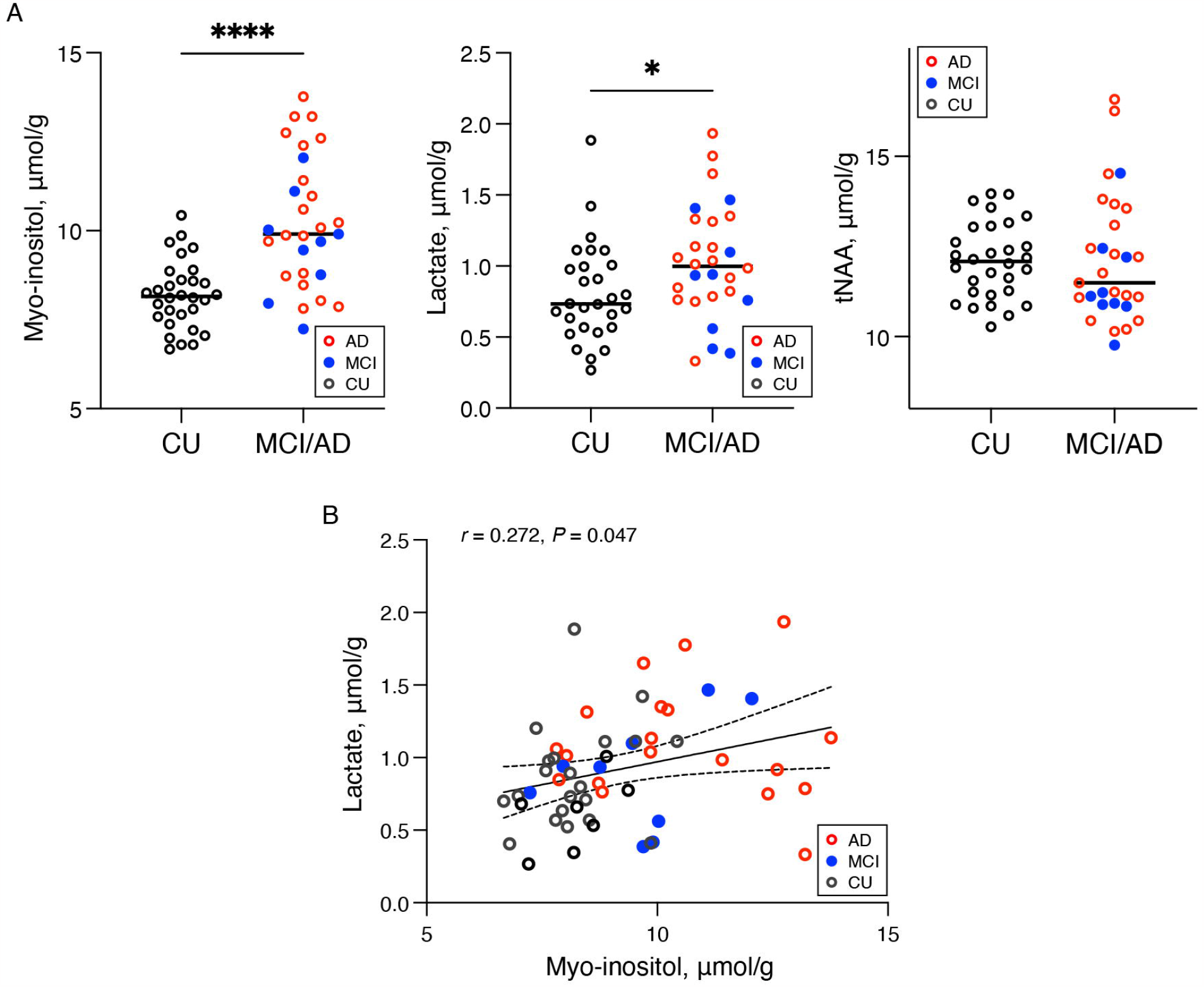
Scatter plots of MRS metabolite levels and association between myo-inositol and lactate levels. (A) Concentrations of myo-inositol, lactate, and tNAA in the PCC in the CU group and AD group. Bars indicate median values. Significance levels: \**p* < 0.05, \*\*\*\**p* < 0.0001. (B) The association between myo-inositol and lactate concentrations in the PCC was tested using partial correlation analysis adjusted for age and sex. A linear regression line with a 95% confidence interval was obtained from the unadjusted analysis. Individual metabolite values are represented by circles in black, blue, and red for the CU, MCI, and AD groups, respectively. AD = Alzheimer disease; CU = cognitively unimpaired controls; MCI = mild cognitive impariment; PCC = posterior cingulate cortex; tNAA = total N-acetyl-aspartate.

### MRS and clinical disease severity

We examined the relationship between MRS metabolites and AD severity using CDR-SOB, which is frequently used as a primary endpoint in clinical trials for AD.^33^ Higher CDR-SOB scores were associated with both high myo-inositol and lactate levels, with a higher correlation with lactate (*r* = 0.452, *p* = 0.018 for myo-inositol, and *r* = 0.528, *p* = 0.006 for lactate) (Figure 3).

**Figure 3.**
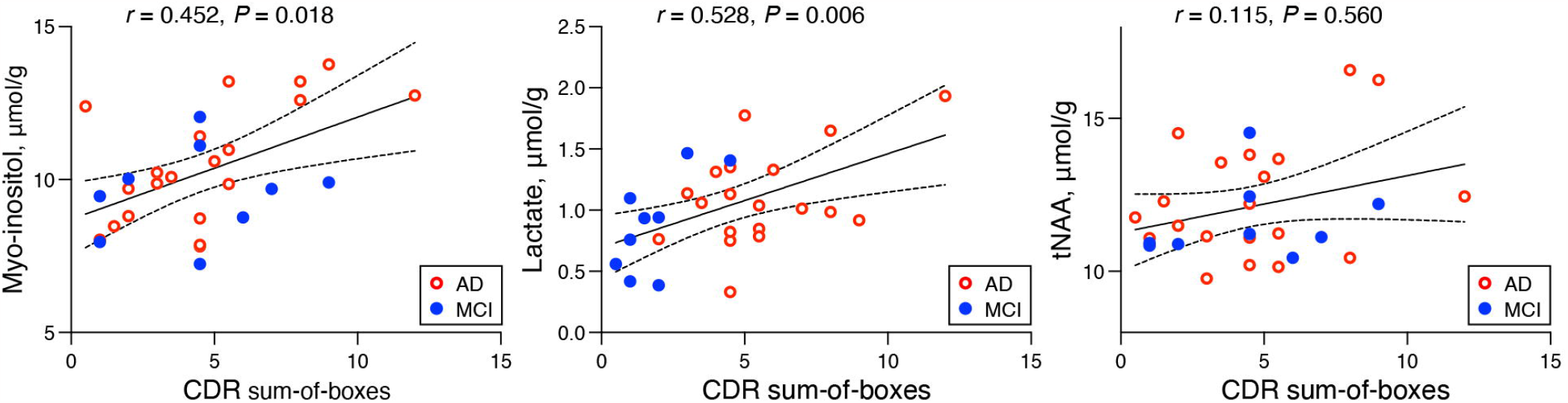
Correlations between MRS metabolites and CDR sum-of-boxes scores. Partial correlation analysis, controlling for age and sex, was conducted to examine the relationship between MRS metabolite levels in the PCC and CDR sum-of-boxes scores. A linear regression line with a 95% confidence interval was derived from the unadjusted analysis. Red and blue circles represent the CU and AD groups, respectively. AD = Alzheimer disease; CDR = Clinical Dementia Rating; CU = cognitively unimpaired controls; MCI = mild cognitive impairment; PCC = posterior cingulate cortex; tNAA = total N-acetyl-aspartate.

### MRS and plasma biomarkers

Finally, we investigated the associations between MRS metabolites and plasma biomarkers. The levels of all plasma biomarkers analyzed in this study were significantly different between the CU and AD groups: the GFAP level increased (*p* < 0.001), Aβ42/40 ratio decreased (*p* < 0.001), and p-tau 181 increased (*p* < 0.001) in the AD group (Figure 4A). A significant positive correlation was found between myo-inositol and plasma GFAP levels (*r* = 0.602, *p* < 0.001). Further, myo-inositol levels correlated significantly with plasma p-tau 181 and showed a tendency to correlate with Aβ42/40 ratio (*r* = 0.457, *p* < 0.001 for p-tau 181; *r* = -0.257, *p* = 0.069 for the Aβ42/40 ratio) (Figure 4B). Lactate levels were not associated with the plasma biomarkers assessed in this study.

**Figure 4.**
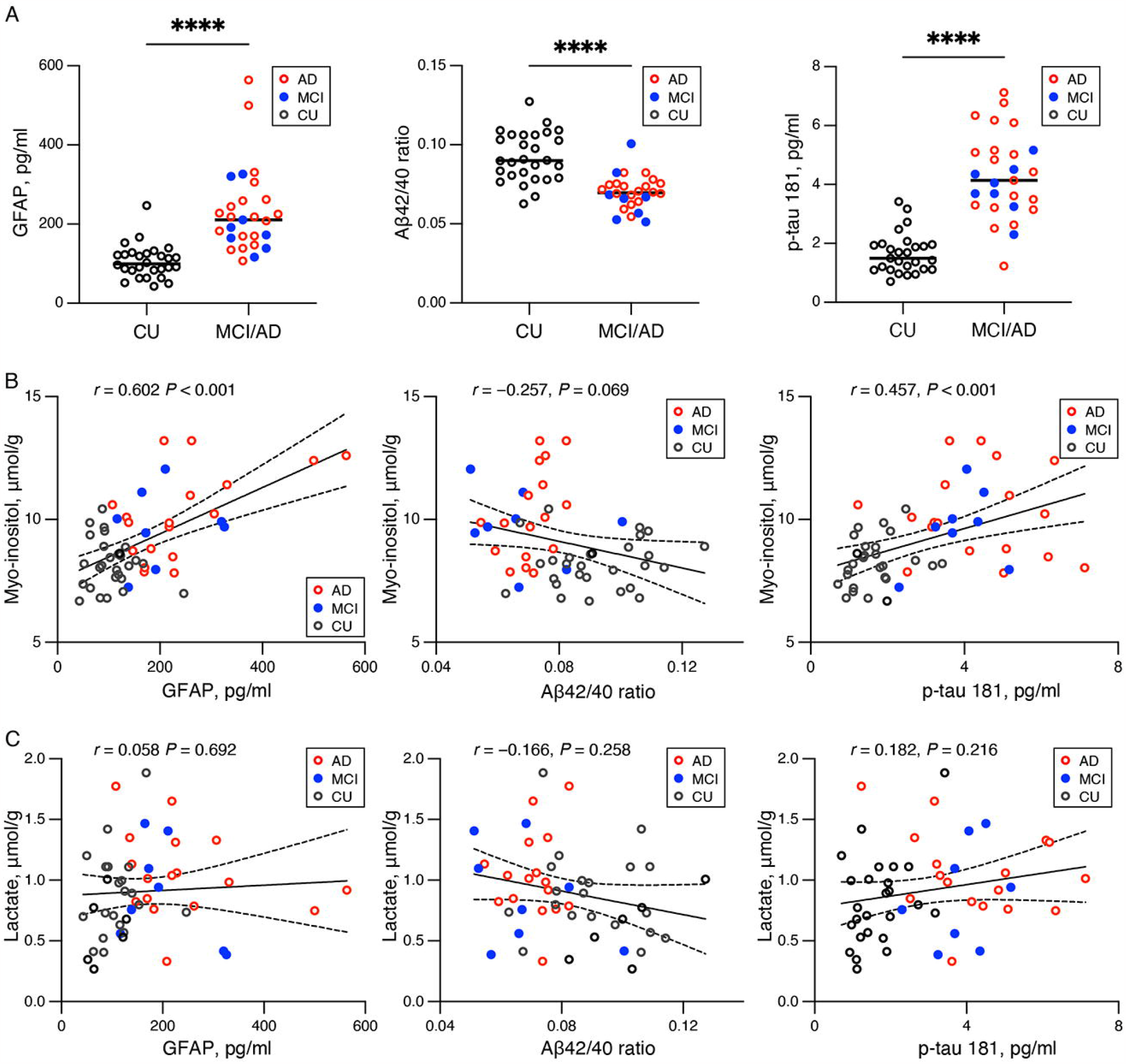
Scatter plots of plasma biomarker levels and association between MRS metabolites and plasma biomarkers. (A) Plasma GFAP, Aβ42/40 ratio, and p-tau 181 levels in the PCC in the CU and AD groups. Bars indicate median values. Significance levels: \*\*\*\**p* < 0.0001. (B) Associations between myo-inositol and lactate concentrations in the PCC and plasma biomarkers were evaluated by partial correlation analysis adjusted for age and sex. A linear regression line with a 95% confidence interval was obtained from the unadjusted analysis. Black, blue, and red circles indicate individual values for the CU, MCI, and AD groups, respectively. AD = Alzheimer disease; Aβ42 = amyloid β 42; Aβ40 = amyloid β 40; Aβ42/40 ratio = ratio Aβ42 to Aβ40; CU = cognitively unimpaired controls; GFAP = glial fibrillary acidic protein; MCI = mild cognitive impairment; p-tau181 = tau phosphorylated at threonine 181.

## DISCUSSION

This study assessed changes in astrocyte activity and brain energy metabolism in patients with AD using MRS. Changes in MRS metabolites were evaluated in the PCC, a region that has been identified as one of the most implicated areas in energy metabolism changes in AD. Our results showed that myo-inositol, an astrocyte marker, and lactate levels were significantly high in the AD group compared with the CU group. To the best of our knowledge, this is the first study that showed a positive association between these metabolites. Additionally, a significant positive correlation was found between myo-inositol and plasma GFAP, two widely used reactive astrocyte biomarkers.

Myo-inositol levels in the PCC were significantly high in the AD group compared with that in the CU group, suggesting astrocyte reactivity. Myo-inositol is mainly found in astrocytes, and high myo-inositol levels were found to be associated with reactive astrogliosis in neurodegenerative diseases including AD.^12^ A correlation between myo-inositol levels and CDR-SOB scores further supports the utility of myo-inositol as a non-invasive biomarker of AD. A novel finding of this study was the positive association between myo-inositol and plasma GFAP levels across the CU and AD groups. Various biomarkers for astrocytes are known, each of which indicates specific aspects of astrocyte reactivity.^3^ Our findings suggest that the two astrocyte markers analyzed in this study may reflect similar changes in astrocytes. Post-mortem studies have found hypertrophy of astrocytes in AD.^34^ Myo-inositol plays a role in maintaining astrocyte cell volume as an osmolyte,^35^ and GFAP, an intermediate filament protein, is a component of the cytoskeleton in astrocytes.^3^ Therefore, the modulation of both myo-inositol and GFAP levels was postulated to be related to morphological changes in reactive astrocytes, resulting in similar biomarker signatures.

Previous studies have shown impaired brain energy metabolism in AD, whereas its molecular and cellular mechanisms remain poorly understood. This study revealed that lactate levels in the PCC were significantly high in the AD group compared with that in the CU group and were associated with AD severity. Despite some attempts to measure lactate concentrations in the CSF of patients with AD, the results have been inconsistent.^36, 37^ The current in vivo measurements of brain lactate levels using MRS provide important data regarding altered lactate concentrations in AD. The increase in brain lactate levels is consistent with previous MRS studies in patients and animal models of AD.^11, 13, 38^ In addition, MRS studies in patients with psychiatric disorders, such as schizophrenia^39^ and bipolar disorder^40^, have reported high lactate levels. This is further supported by a recent study using various mouse models of neuropsychiatric disorders.^10^ Lactate accumulations can negatively affect neuronal energy homeostasis and brain activity. Furthermore, high lactate levels in the brain can disrupt normal cellular processes and impair physiological function by decreasing the pH.^41^ These findings indicate that high lactate levels are associated with the pathogenesis of not only AD but also other neuropsychiatric disorders.

The finding of a positive correlation between myo-inositol and lactate levels in this study suggests a link between reactive astrocytes and altered brain energy metabolism. High lactate levels in the brain can be caused by increased production and/or decreased clearance/utilization. Anaerobic metabolism, often resulting from mitochondrial dysfunction and hypoxia/ischemia, has long been associated with increased lactate production. Mitochondrial dysfunction, primarily in neurons, has been widely documented in AD studies.^42^ Impaired lactate clearance, as indicated by the impaired blood– brain barrier, was also proposed as a contributing factor.^43^ The correlation between myo-inositol and lactate levels in this study suggests another mechanism underlying high lactate levels in the brain. It has been suggested that astrocytes are the primary source of lactate production in the brain, shuttling lactate via monocarboxylate transporters (MCTs) to neurons as an energy source, a process known as ANLS.^5, 8^ Accordingly, an impaired ANLS system may contribute to lactate level dysregulation. Studies revealed that reactive astrocytes demonstrate dynamic increases in calcium signaling,^44^ and an in vitro study showed that these signals act as key triggers for the augmented glucose uptake and aerobic glycolysis in astrocytes that results in high lactate levels.^45^ Therefore, aberrant calcium signaling in reactive astrocytes may lead to excessive lactate production. Additionally, a previous study reported a reduction in MCTs expression in an AD mouse model,^46^ suggesting that lactate is not efficiently released from astrocytes to neurons, leading to a disruption in energy utilization and surplus lactate levels. However, it should be noted that there is a body of literature questioning the ANLS model.^47^ Thus, further research is necessary to fully comprehend the underlying mechanisms and evaluate whether targeting astrocytic metabolism may be a potential therapeutic approach for AD.

In the present study, a significant correlation was found between myo-inositol levels and plasma p-tau 181 levels, with a trend towards a correlation with the Aβ42/40 ratio, although not statistically significant. This finding is in line with those of previous studies demonstrating an association between reactive astrogliosis and tau and Aβ pathologies in the brain.^48^

A previous MRS study reported that the administration of anti-Aβ antibody to APP-PS1 mice attenuated the increases in myo-inositol, indicating that high myo-inositol levels may be attributed to Aβ pathology.^49^ However, research investigating the relationship between high myo-inositol levels and tau pathology is scarce. The concurrent presence of Aβ and tau in the brains of patients with AD makes it difficult to discern the individual effects of each pathology. Further studies using tau model mice are necessary to determine the influence of tau pathology on myo-inositol levels.

This study has several limitations. Given the challenges in measuring lactate because of its low concentration, previous lactate measurements in MRS have relied on high-field MRI such as 7-T or 3-T MRI with specific methods.^13, 39, 40^ In this study, lactate was measured using 3-T MRI with an advanced MRS protocol, SPECIAL sequence. The mean CRLB for lactate in this study (24.9%) was comparable to that of a previous study using 7-T MRI to measure lactate (20.6% for the control participants),^39^ and the correlation between lactate and disease severity further supports data reliability. Second, lactate levels were evaluated in a steady state, and the dynamics of lactate metabolism was not assessed. Third, MRS data were acquired from a single VOI in the PCC, and different brain regions may show different patterns of metabolic changes in AD.^50^

## Conclusions

In this study, in vivo assessment using MRS was employed to highlight changes in brain energy metabolism associated with astrocyte reactivity, offering novel insights into the role of reactive astrocytes in AD pathogenesis. High lactate levels detected by MRS may be a valuable biomarker for assessing altered brain energy metabolism. Myo-inositol and plasma GFAP, two astrocyte markers, may reflect similar changes in astrocytes.

## Data Availability

All data produced in the present study are available upon reasonable request to the authors

## Acknowledgements

The authors thank all patients and their caregivers for participation in this study, and clinical research coordinators, PET and MRI operators, animal care technicians, radiochemists, and research ethics advisers at QST for their assistance to the current projects. We thank APRINOIA Therapeutics for kindly sharing the precursor of [^18^F]florzolotau. The authors acknowledge the support of Dr. Shigeki Hirano at the Chiba University, Dr. Yasumasa Yoshiyama at the Inage Neurology and Memory Clinic, and Dr. Shunichiro Shinagawa at the Jikei University School of Medicine, for patient recruitment.

## Author Contributions

K.H., K.M., M.O., T.T., M.H., and Y.T. contributed to the conception and design of the study. K.H., K.M., K.T., H.E., H.T., N.K., H.S., K.T., T.O., J.N., K.K., M.-R.Z., H.Sd., T.Y., and Y.T. contributed to data acquisition and analysis. K.H., K.M., M.O., and Y.T. contributed to drafting the text or preparing the figures.

